# Entomological surveillance during a major CHIKV outbreak in northwestern São Paulo: insights from São José do Rio Preto

**DOI:** 10.1101/2024.12.04.24318429

**Authors:** Cecília Artico Banho, Maisa Carla Pereira Parra, Olivia Borghi Nascimento, Gabriel Pires Magnani, Maria Vitoria Moraes Ferreira, Ana Paula Lemos, Beatriz de Carvalho Marques, Marini Lino Brancini, Livia Sacchetto, Andreia Francesli Negri, Regiane Maria Tironi Menezes, Juliana Telles de Deus, Cassia Fernanda Estofolete, Nikos Vasilakis, Maurício Lacerda Nogueira

## Abstract

**Background:** Brazil is considered an epicenter for emerging and re-emerging arboviruses that significantly impact public health. The mid-sized city of São José do Rio Preto (SJdRP) in northwestern São Paulo state is considered hyperendemic for arboviral diseases, with case numbers climbing each year. Only 45 cases of chikungunya (CHIKV) were reported in the city from 2015 to 2022, indicating cryptic circulation of this virus, but cases in the state increased notably in 2023. This study investigates the use of active entomological surveillance to detect new arbovirus introductions in specific areas like SJdRP.

**Methodology/Principal findings:** We used molecular testing to investigate the presence of CHIKV in adult culicids collected monthly from various neighborhoods in SJdRP. Positive samples underwent whole-genome sequencing and phylogenetic analysis. Entomological surveillance successfully detected the early spread of CHIKV across SJdRP, revealing an infection rate of 6.67%, with the well-established vectors *Aedes aegypti* and *Ae. albopictus* as well as *Culex* sp. carrying the virus. The vector positivity rate increased from December 2023 to April 2024, which correlates with rising numbers of chikungunya fever cases reported in SJdRP during the same period. The resurgence of CHIKV in this region is attributed to several introduction events, mainly from the Southeast and North of Brazil, which facilitated establishment of the virus within the highly dense vector population and led to extensive spread and, in turn, a major CHIKV epidemic in this geographical area.

**Conclusions/significance:** Extensive circulation of CHIKV was documented within the human and vector population, marking the onset of the first major CHIKV epidemic in SJdRP and neighboring cities. Because multiple arboviruses co-circulate in several locations in Brazil, entomological surveillance, along with ongoing monitoring of patient samples, is a key to help health authorities to implement more effective measures to interrupt transmission cycles and mitigate new epidemic waves.

**Author summary:** The city of São José do Rio Preto (SJdRP) in northwestern São Paulo state is an endemic area for dengue virus (DENV), but cases of chikungunya virus (CHIKV) were also reported between January and September 2023. Since overlapping symptoms between these acute febrile diseases can complicate differential diagnosis, the increase in CHIKV cases in DENV-endemic regions is concerning. Entomological surveillance is a useful strategy for accurate and early detection of arboviruses, making it possible to identify emerging or increased arbovirus activity, predict potential outbreaks, and support effective control measures, thus reducing impacts on public health. Through entomological surveillance we were able to detect the spread of CHIKV in SJdRP, revealing a high infection rate in the vector population. Our findings also suggest that the virus spread widely throughout the local mosquito population, potentially via vertical or sexual transmission, which may have contributed to sustained transmission during unfavorable conditions or inter-epidemic periods. We also observed a monthly increase in the vector population’s positivity rate which correlates with a rise in CHIKV cases in the city and the first CHIKV outbreak in this area.

## Introduction

Chikungunya virus (CHIKV) (*Togaviridae, Alphavirus*) is one of the main arthropod-borne viruses transmitted by *Aedes aegypti* and *Aedes albopictus* [1]. It causes a substantial impact on global public health, especially in tropical and subtropical areas where competent vectors are widely distributed [2].

The presence of CHIKV in the Americas was first detected in 2013 [3], and the introduction of the Asian and East/Central/South-African (ECSA) genotypes was subsequently reported in the North and Northeast regions of Brazil, respectively [4]. The ECSA genotype was first identified in September 2014 in the state of Bahia, and it rapidly spread throughout the country, causing annual epidemic waves of chikungunya fever (CHIKF), particularly in the Northeast and Southeast of the country [4]. Between 2017 and 2024 Brazil reported 936,854 confirmed cases, 205,718 of these registered during January–November 2024 [5]. Within Brazil, an alarming number of confirmed cases are appearing in the states in the Southeast region, which accounted for 74.7% of all CHIKF cases reported in 2024 [5]. The rise of CHIKV infections in Southeast Brazil is driven by multiple factors that include 1) high infestation rates of *Ae. aegypti* and *Ae. albopictus*, especially in São Paulo state [6]; 2) large, populous metropolitan areas with inadequate sanitation that allows *Aedes* mosquitoes to proliferate; 3) climate change (since the rise in average temperatures during drier and cooler seasons creates favorable conditions for established mosquito populations to breed for longer periods of time) [7]; and 4) a population susceptible to CHIKV infection, as indicated by the low anti-CHIKV IgG seroprevalence reported in all major cities in São Paulo state [8,9].

This is a concerning scenario, as CHIKV currently cocirculates with other high-incidence arboviruses in several Southeastern cities [7] such as dengue virus (DENV) serotypes 1, 2, and 3 [10–12]. Moreover, the growth of CHIKF in DENV-endemic areas presents a challenge for differential diagnosis, since the symptoms of these acute febrile diseases overlap. This situation highlights the need for continuous monitoring of circulating arboviruses, since CHIKV can cause long-lasting sequelae such as debilitating arthritis and arthralgia or more severe outcomes like neurological disorders [13].

Entomological surveillance coupled with differential diagnosis is essential to improve the accuracy and early detection of arboviruses in a specific region. This dual approach can facilitate early identification of arbovirus activity, make it possible to predict outbreaks, and guide the development of more effective control measures to reduce their impact on human health. A recent study in São José do Rio Preto (SJdRP), a medium-sized city in northwestern São Paulo state that is classified as hyperendemic for dengue [9], revealed low circulation of CHIKV from 2015 to 2019. This study warned of potential outbreaks of this disease in the future, due to the presence of competent vectors and a substantial immunologically naïve CHIKV population [9]. Along similar lines, considering the recent rise in CHIKF cases in São Paulo state [5] we investigated the use of active entomological surveillance for early detection of new arbovirus introductions that could contribute to a new epidemic wave, and also used this data to trace the spread of this virus in the region.

## Material and methods

### Study area

São José do Rio Preto (SJdRP) is a city in northwestern São Paulo state with an estimated 480,393 inhabitants, 94% in urban areas and 6% in rural settings [14]; average annual temperature and rainfall are 27 °C and 139 mm, respectively [15]. It is the largest municipality in the northwestern region of the state and the headquarters for the XV Regional Health District (RHD-XV), which comprises 101 adjacent municipalities. The city is considered hyperendemic for arboviral diseases, and a high number of arboviral infections are recorded here, primarily DENV [16].

### Recruitment and collection points

Because of this hyperendemic status, in SJdRP we established a sentinel program for early detection of emergent or highly active arboviruses within vectors in urban areas. First, we conducted an epidemiological survey in partnership with the city department of health to determine the neighborhoods with the most reported arbovirus cases and/or vector infestations over the past five years. Six neighborhoods were selected for this study and arranged into a north/south and east/west grid of the municipality. For collection points, we prioritized locations that shielded traps from direct sunlight and rainfall near areas with accumulated trash or recyclable materials and vacant lots with vegetation. Because we needed to obtain consent from residents for monthly visits, the number of collection points could not be standardized across the six selected neighborhoods; 46 residences were recruited as collection sites. Fieldwork began in October 2023 with recruitment and obtaining consent to authorize installation of mosquito traps in intra- and/or peridomestic areas. Mosquitoes were collected from the traps every month, and verbal consent for the field agents to enter the properties was again requested from residents during all visits. Adult mosquitoes were trapped using BG-Sentinel traps (Biogents, Germany) installed outdoors in shaded sites and/or near vegetation and maintained at the collection points for 24 hours. After this period, the specimens were transferred to appropriate containers and transported to the virology laboratory at our institution r species-level identification using taxonomic keys [17,18]. After identification, the collected specimens were stored individually in 1.5 ml polypropylene tubes in a -80°C freezer until subsequent analyses.

This study was approved by our institutional ethics review board. All data were analyzed anonymously, ensuring total confidentiality for all participants.

### Geoprocessing

For spatial analysis, a database was created in Microsoft Excel with information on the collected mosquitoes (collection location, identification number, and results). Shapefiles of the areas were provided by the SJdRP city government and obtained from the Brazilian Institute of Geography and Statistics (IBGE) [19]. Maps were created with R v.4.0.114 software [20], using the sf v. 1.0-16 [21,22], and ggplot2 [23] packages.

Epidemiological data for confirmed cases of CHIKF during 2017–2024 for all Brazilian states were obtained from the Brazilian Ministry of Health [5]; Chikungunya incidence was calculated per 100,000 inhabitants based on the estimated populations of Brazilian states from 2017 to 2024 as reported by the Brazilian Institute of Geography and Statistics in its SIDRA database [24], available at https://sidra.ibge.gov.br/tabela/7358, accessed on August 15, 2024. Case numbers for CHIKF reported in SJdRP were obtained from the city health department [25].

### Arbovirus detection

The adult mosquitoes collected each month were first macerated in 400 μL of 1X PBS solution with a homogenizer bead, using the L-BEADER mechanical cell disruptor (Loccus, Brazil) in three cycles of 30 seconds at 3000 rpm. The samples were then centrifuged at 7,500 rpm for ten minutes and the mosquito macerates were used for viral RNA extraction, according to Machado et al. [26]. Next, molecular analyses were conducted to detect the presence of CHIKV. One-step real-time polymerase chain reaction (RT-qPCR) was performed using the GoTaq Probe 1-Step RT-qPCR system (Promega, Madison, USA) along with TaqMan fluorescent primers and probes specific to CHIKV obtained from Lanciotti et al. [27]. The reactions were conducted with a QuantStudio 3 Real-Time PCR System (Thermo Fisher Scientific, MA, USA), and cycle threshold (Ct) values below 38 results were considered positive. All analyses were performed in linear and multi-component mode, adjusting the baseline to the Ct of the negative control to eliminate possible reagent interference.

### CHIKV whole-genome sequencing

After molecular screening for CHIKV in the mosquitoes, positive samples underwent whole-genome sequencing. Library construction and complete genome sequencing were performed using next-generation sequencing (NGS); cDNA synthesis, genome amplification, and library preparation were carried out according to the instructions provided for the Illumina CovidSeq Test (Illumina, San Diego, CA, USA), but adapted by replacing the SARS-CoV-2 primer pools with CHIKV-specific primer pools designed by the Brazil-UK Centre for Arbovirus Discovery, Diagnosis, Genomics, and Epidemiology (CADDE, available at: https://www.caddecentre.org/pt/protocols-pt/). Library quantification was performed using the Qubit dsDNA HS Assay on a Qubit 2.0 device (Invitrogen, Waltham, MA, USA). Quality control for the libraries was verified with a TapeStation 4150 system and High Sensitivity D1000 ScreenTape kit (Agilent Technologies, Santa Clara, CA, USA). Sequencing was done with a MiSeq Reagent kit v3 (2 x 150 cycles) (Illumina, San Diego, CA, USA), and the NextSeqTM PhiX Control kit was used as a normalization sample with the libraries sequenced on the MiSeq system (Illumina, San Diego, CA, USA).

### Serum samples

In order to link detection of arbovirus-positive mosquitoes in SJdRP with human cases and better characterize the CHIKV genotype circulating in northwestern São Paulo state, we included 83 serum samples from patients with a positive diagnosis of CHIKF. These human samples were collected at a tertiary hospital in SJdRP that serves patients from neighboring cities within the regional health district. The samples were obtained during January–May 2024 from hospitalized and non-hospitalized individuals residing in 13 different municipalities within the RHD-XV and two cities in the adjacent state of Minas Gerais (Frutal and Fronteira) which border São Paulo and are less than 115 kilometers from SJdRP. These analyses utilized retrospective samples collected for routine diagnosis, and the need for informed consent was waived by our institutional review board.

Total RNA extraction was performed on all serum samples using a QIAamp Viral RNA Mini Kit (QIAGEN, Hilden, Germany), following the manufacturer’s instructions. Next, RT-qPCRs to detect CHIKV and whole-genome sequencing were performed as previously described.

### Genome assembly and phylogenetic analyses

The quality of raw reads was assessed using the FastQC v. 0.11.4 program [28], and Cutadapt v. 4.6 software [29] was used to remove low-quality reads (Phred score >30) shorter than 50 base pairs (bp), as well as duplicate sequences, adapters, and primers used during library construction. Clean reads were then mapped against the genomes of their respective hosts (GCA_002204515.1, GCA_001444175.1, and GCA_015732765.1, available at: https://vectorbase.org/vectorbase/app/search/organism/GenomeDataTypes/result) using Bowtie2 v. 2.5.25 [30]. Reads not mapped to the host genome were filtered using SAMtools v1.10 [31] and mapped against the CHIKV reference genome (NC_004162.2) using BWA mem v. 0.7.17 software [32] and SAMtools v. 1.10[31] for read sorting and indexing. After post-processing steps, the assembled genomes were recovered and using iVar v. 1.3.19 [33].

Next, all the generated consensus sequences were analyzed using the Genome Detective virus typing tool [34] for genotype classification. Phylogenetic analyses were also performed to confirm the genotype of the circulating virus and divergence from viruses sequenced from other Brazilian regions. To do so, the assembled genomes were aligned with a dataset containing CHIKV sequences available in the EpiArbo-GISAID database [35] and the GenBank NCBI database [36], using MAFFT v. 7.27110 [37] and edited with AliView v. 1.28 [38]. Maximum likelihood (ML) trees were reconstructed using IQ-TREE v. 2.0.3.711 software [39], with the best nucleotide substitution model inferred according to the Bayesian information criterion (BIC) by ModelFinder [40]. Branch reliability was tested using a combination of the ultrafast bootstrap approximation approach (UFBoot) [41] and SH-like approximate likelihood ratio test (SH-aLRT) [42] with 10,000 replicates each, respectively. Phylogenetic trees were visualized and edited using R v.4.0.114 software [20] and the ggtree package [43].

To investigate the temporal signal from the ML trees, we regressed root-to-tip genetic distances against sample collection dates using the TempEst tool v. 1.5.1[44], considering a correlation coefficient of >0.4 to accept temporal structure. Next, the generated phylogenies were subjected to TreeTime v. 0.9.3 [45] to convert the raw ML trees into time-scaled trees, as described by Banho et al. [46]. Finally, we used the time-scaled tree topologies to infer the number of viral exchange events between the five Brazilian regions, the cities within the RHD-XV and SJdRP using TreeTime mugration v. 0.9.3 [45], and by mapping the locations to tips and internal nodes from the annotated tree topology we were able to estimate the number of virus importations and exportations among regions/cities.

## Results

### CHIKV prevalence in Brazil and in SJdRP, 2017–2024

Several waves of infection have been observed since the introduction of CHIKV into Brazil (Fig. 1A). According to the National Reportable Disease Information System (SINAN) [5], a total of 916,115 confirmed cases of CHIKF were reported in all Brazilian states from January 2017 to July 2024 ( S1 Table). During this same period eight peaks of infections were identified, predominantly in April of each year. The North and Southeast regions of the country accounted for most of these reported cases (Fig. 1A), but the incidence rates per 100,000 inhabitants clearly show that in 2017 the highest incidence of CHIKF was observed in the North and Northeast, particularly in the states of Roraima and Ceará, which reported 723 and 1,196 cases per 100,000 inhabitants, respectively. In 2018, the states with the highest incidence shifted to Pará, Mato Grosso, Rio de Janeiro, and Minas Gerais, in the North, Midwest, and Southeast regions of the country, respectively (Fig. 1B, S2 Table). During 2020 and 2021, the highest incidence of CHIKF cases was concentrated in the Northeast, but from 2022 an increase was also observed in the Southeast as well as certain states in the Midwest and North (Fig. 1B and S2 Table).

**Fig 1.**
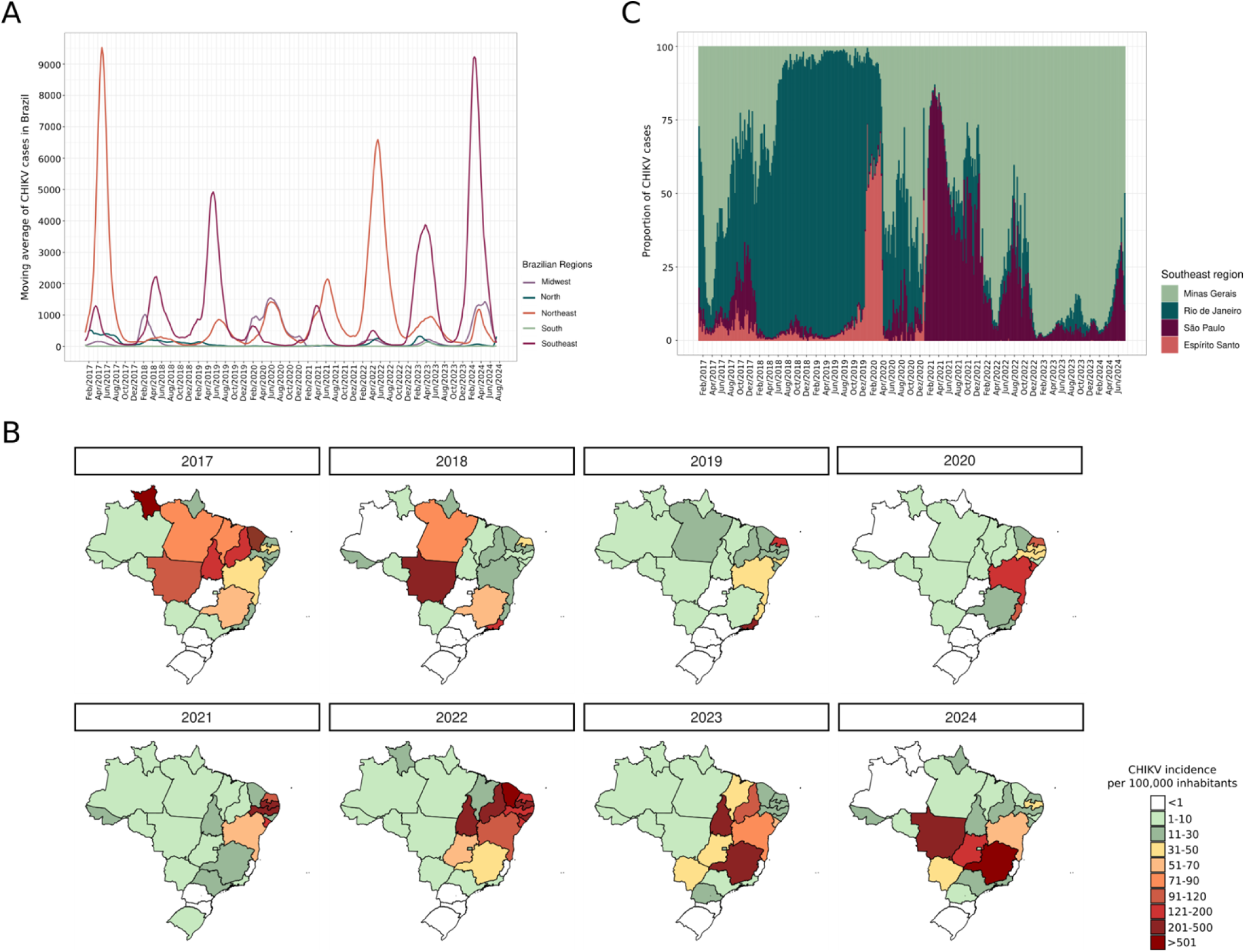
Laboratory confirmed cases of Chikungunya fever in Brazil. A) Moving average of confirmed CHIKF cases across all five Brazilian regions from 2017 to 2024, highlighting several peaks of infection. B) Incidence rate of CHIKF per 100,000 inhabitants in all Brazilian states from 2017 to 2024, with color scale representing variation in incidence rates. C) Proportion of reported CHIKF cases in the Southeastern states of Brazil from 2017 to 2024.

Among the Southeastern states, Minas Gerais has been the most severely impacted by CHIKV infections. The number of reported CHIKF cases soared from 2022 to reach 258,286 confirmed cases in 2024 (Fig. 1C). Similarly, the number of confirmed CHIKF cases in São Paulo also rose from 2022.

### Entomological surveillance as a tool for early detection of arboviruses

Considering the significant increase in CHIKF cases in the state of São Paulo, which could potentially lead to outbreaks in cities already severely affected by other arboviral diseases like dengue, a mosquito surveillance program was established in SJdRP from October 2023. The city is considered hyperendemic for arboviruses, with a concerning number of confirmed dengue cases and three different serotypes (DENV-1-3) co-circulating in late 2023 and 2024 [16]. In early 2023 some CHIKV infections were reported in the city, with 17 confirmed cases from January to September 2023 [25].

Between October 2023 and July 2024, our entomological surveillance system collected and identified a total of 1,183 culicid specimens: *Ae. aegypti* (n=744/1,183, 62.9%), *Ae. albopictus* (n=11/1,183, 0.92%), *Aedes sp*. (n=1/1,183, 0.08%), and *Culex sp*. (n=427/1,183, 36.0%).

Specimens of *Aedes* and *Culex* were collected throughout the study period, but a distinct seasonal abundance pattern was observed. *Aedes* mosquitoes were more prevalent from October 2023 to April 2024, peaking in February 2024 when 178 (23.5%) *Aedes* specimens were collected. In contrast, the number of *Culex* mosquitoes collected increased from May 2024 while the abundance of *Aedes* specimens decreased (Fig. 2A, S3 Table).

**Fig 2.**
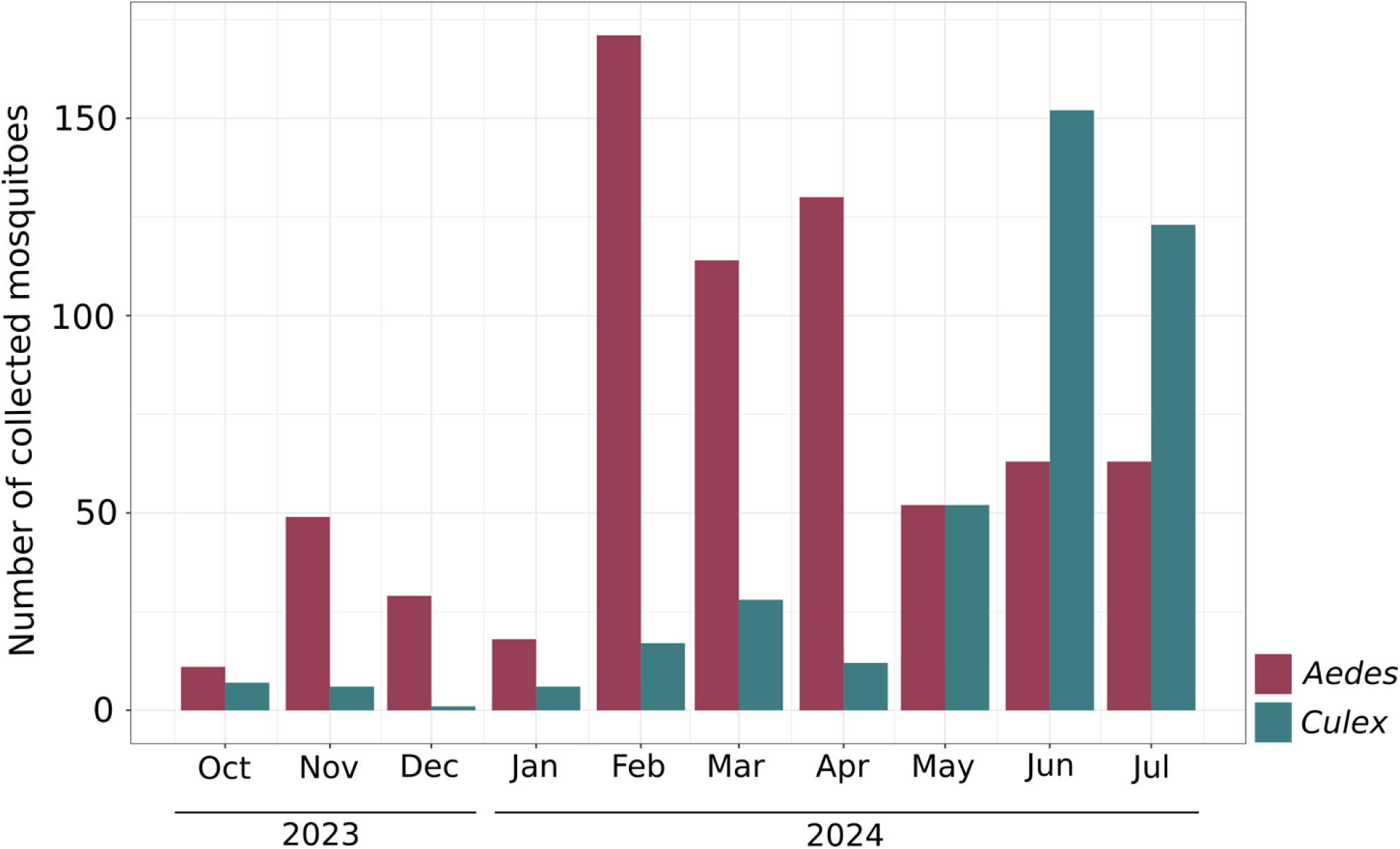
Vector surveillance. Number of mosquitoes collected per month and genus, October 2023–July 2024.

Next, all the collected specimens underwent molecular testing to detect the presence of CHIKV: 79 mosquitoes were positive for CHIKV, with an overall positivity rate of 6.67%. The positive samples were 26 *Ae. aegypti* females (33%), 27 *Ae. aegypti* males (34%), two *Ae. albopictus* females (2.5%), 17 *Culex sp.* females (21.5%), and seven *Culex sp*. males (8.8%). Notably, only five collected females that tested positive for CHIKV were engorged (one *Ae. aegypti* and four *Culex* sp.) (S3 Table). Considering monthly positivity, the most infected mosquitoes were observed in May 2024 (n= 17/104, 25.96%). Similar results were observed for only *Aedes* mosquitoes, which are widely known as competent vectors for CHIKV (Fig. 3A, S3 Table).

**Figure 3.**
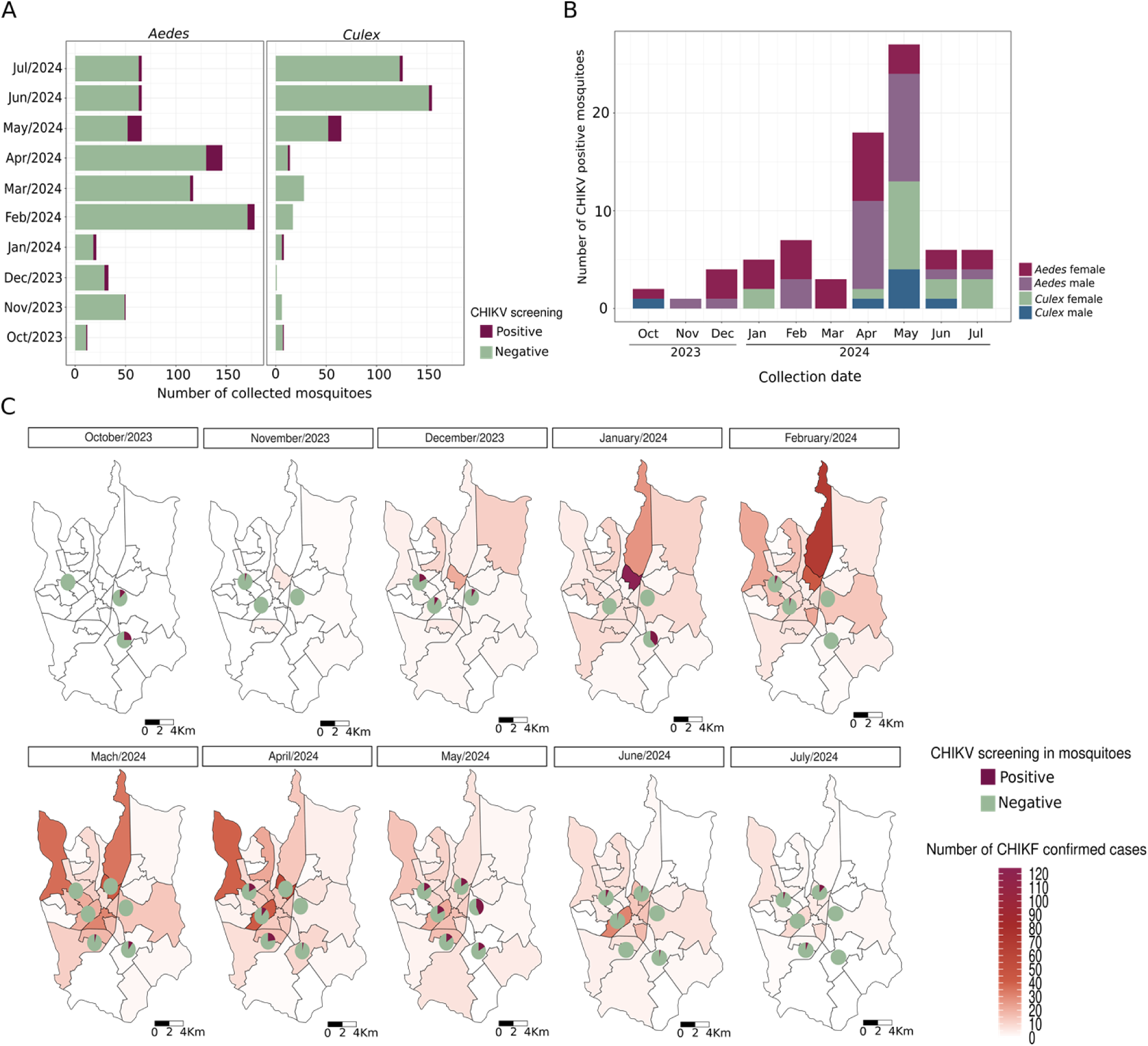
Entomological Surveillance. A) Number of *Aedes* and *Culex* mosquitoes testing negative and positive for CHIKV, October 2023–July 2024. B) Number of *Aedes* and *Culex* mosquitoes that tested positive for CHIKV each month, by sex. C) Temporal analysis of confirmed CHIKF cases in SJdRP, along with the proportion of mosquitoes collected in each neighborhood that tested either negative or positive for CHIKV.

It is important to note that male mosquitoes testing positive for CHIKV were collected in nearly every sampled month (Fig. 3B), suggesting that the virus was widely disseminated throughout the natural mosquito population, potentially via vertical or sexual transmission. This observation was corroborated by a male *Culex* sp. (a species not known to be a vector for CHIKV) that tested positive for the virus in October 2023, a month when no CHIKF cases were reported in the city (Figs. 3C and 4). Moreover, positive mosquitoes were found throughout the study period in various sampled neighborhoods, highlighting extensive virus dissemination across SJdRP (Fig. 3C). Considering positivity by species and sex, most of the positive specimens between December 2023 and April 2024 were *Ae. aegypti* females, with rates ranging from 3.9 to 23% (Fig. 3B). This finding is in line with the rising number of CHIKF cases reported in the city during the same period, with a infections peaking in May 2024 (Figs. 3C and 4).

**Fig 4.**
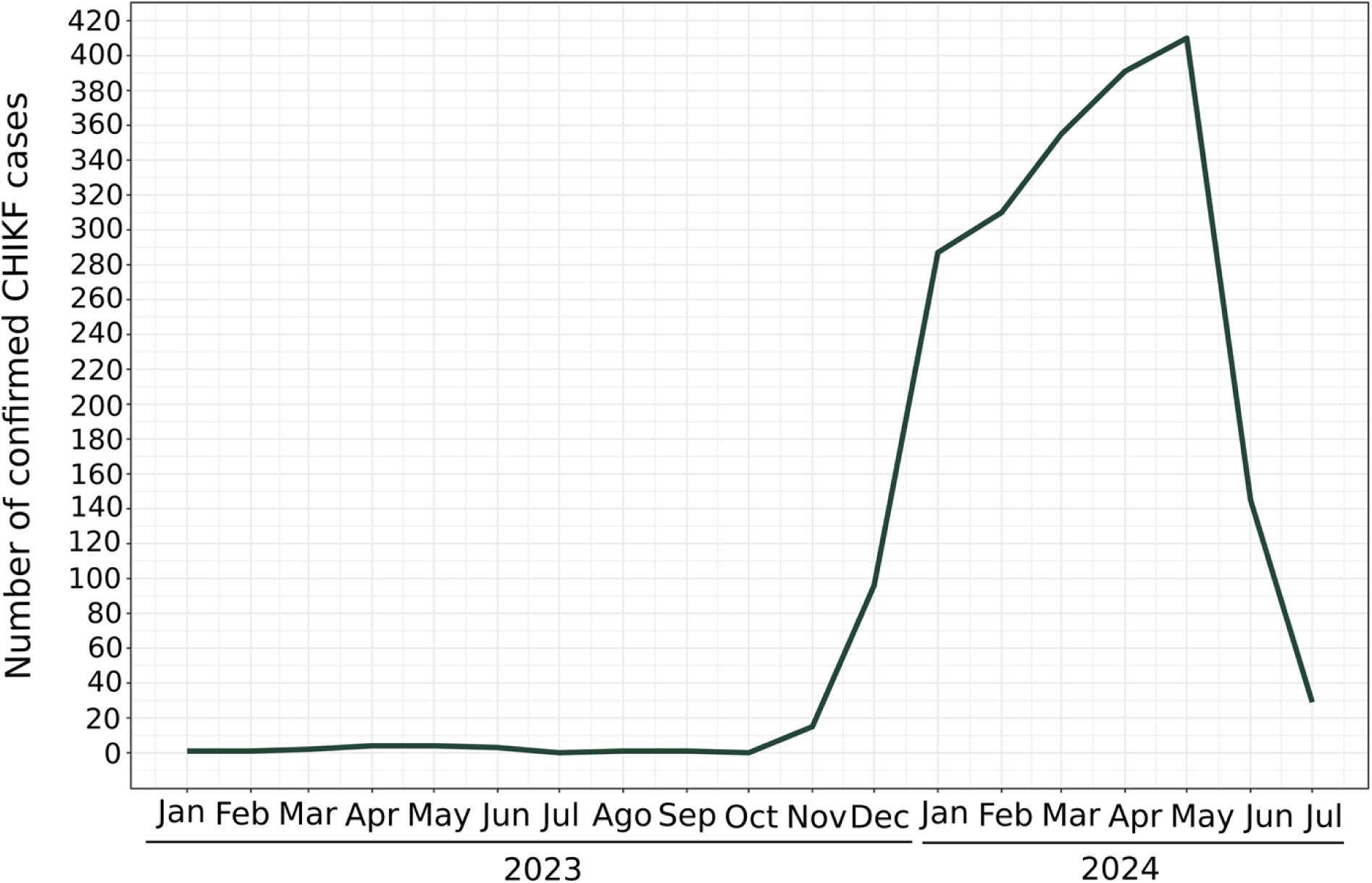
Chikungunya virus surveillance in human samples. Number of confirmed CHIKF cases in residents of SJdRP, by month.

### Resurgence of CHIKV in northwestern São Paulo state

Using next-generation sequencing of CHIKV-positive samples, we successfully recovered the complete CHIKV genome from 66 mosquito specimens (23 *Ae. aegypti* females, 19 *Ae. aegypti* males, two *Ae. albopictus* females, 15 *Culex* sp. females, and seven *Culex* sp. males) as well as from all the human samples. Despite the high cycle threshold values obtained in the mosquito samples, which ranged from 29.9 to 38.0 (S1 Fig, S4 Table), genome coverage exceeded 70%.

The phylogenetic dataset comprises Brazilian sequences collected between 2014 and 2024 (S5 Table). Temporal analysis via root-to-tip genetic distance regression revealed that the dataset exhibits a suitable temporal signal (R² = 0.44 and correlation coefficient = 0.66) that permits reconstruction of a time-scaled phylogenetic tree (Fig. 5).

**Fig 5.**
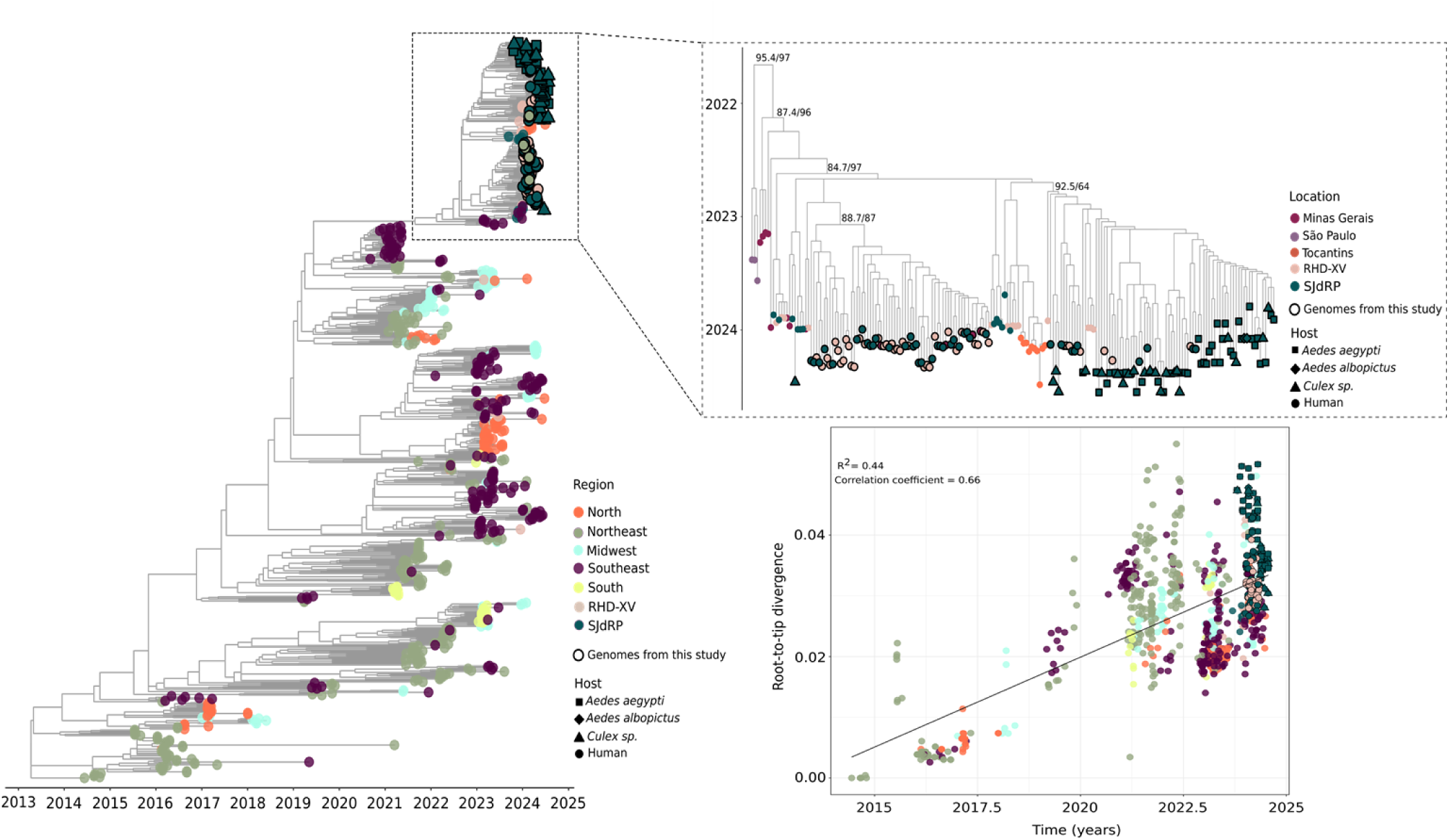
Maximum likelihood tree for CHIKV based on complete genome sequences from SJdRP, other cities within the RHD-XV and all Brazilian regions. Time-stamped phylogenetic tree reconstructed using 782 Brazilian complete genomes (149 from this study) from all Brazilian regions (S5 Table), highlighting the clade formed by CHIKV sequences obtained from *Aedes*, *Culex* and human samples generated in this study, which are grouped with sequences from Minas Gerais, São Paulo and Tocantins states. Linear regression of root-to-tip genetic distance of CHIKV versus sampling date. Colors represent different regions or locations, and tip shapes represent different hosts. Genome sequences from mosquitoes and human samples obtained in this study are highlighted with black borders.

Our maximum likelihood phylogenetic tree indicated that all sequences from this study formed a well-supported clade (UFBoot/alrt = 95.4/97), along with sequences from the Southeast and North regions of Brazil. The CHIKV genomes derived from human and mosquito samples were predominantly divided into two monophyletic groups, exhibiting high branch support (Fig. 5). One group comprised exclusively human sequences from several locations including SJdRP, cities within the RHD-XV region, and municipalities in Minas Gerais bordering the state of São Paulo. The second clade consisted of CHIKV sequences from different mosquito hosts (*Ae. aegypti*, *Ae. albopictus* and *Culex* sp.) as well as human samples, primarily collected from SJdRP and with less representation from RHD-XV municipalities. Importantly, RHD-XV sequences within both clades were interspersed with those from SJdRP, suggesting significant viral exchange between these locations (Fig. 5).

Notably, no distinct clusters were formed according to host: sequences derived from *Aedes* and *Culex* samples were grouped together with high branch support, indicating considerable genetic similarity. Our root-to-tip genetic distance analysis supported these findings, demonstrating that these sequences were clustered with high similarity (Fig. 5). Further analyses revealed that all sequences from SJdRP (collected in this study and also retrieved from the GISAID database) were closely related to CHIKV genomes from Minas Gerais in Brazil’s Southeast region as well as genomes from Tocantins in the North, all collected between 2022 and 2023. Interestingly, examination of the nodes in the ML tree that correspond to the most recent common ancestor (TMRCA) of the sequences obtained in this study shows that they date back to late 2022 and early 2023 (Fig. 5), correlating with the incidence rates of CHIKF cases reported in 2022 and 2023, particularly highlighting the states of Tocantins and Minas Gerais (Fig. 1B). This suggests that both states may have contributed to the recent introduction and increased circulation of CHIKV in the state of São Paulo.

Our migration pattern analysis also revealed a substantial amount of viral exchange, primarily between SJdRP and other municipalities within the RHD-XV region (Fig. 6A), reinforcing the findings from our phylogenetic analysis. Furthermore, our results indicate that Southeastern states are a significant source of CHIKV exportation to other regions of Brazil; this may be associated with the recent wave of infections affecting this region, as well as the considerable number of sequences from São Paulo and Minas Gerais collected from 2022 onward that are included in our dataset (Fig. 6A, S5 Table).

**Fig 6.**
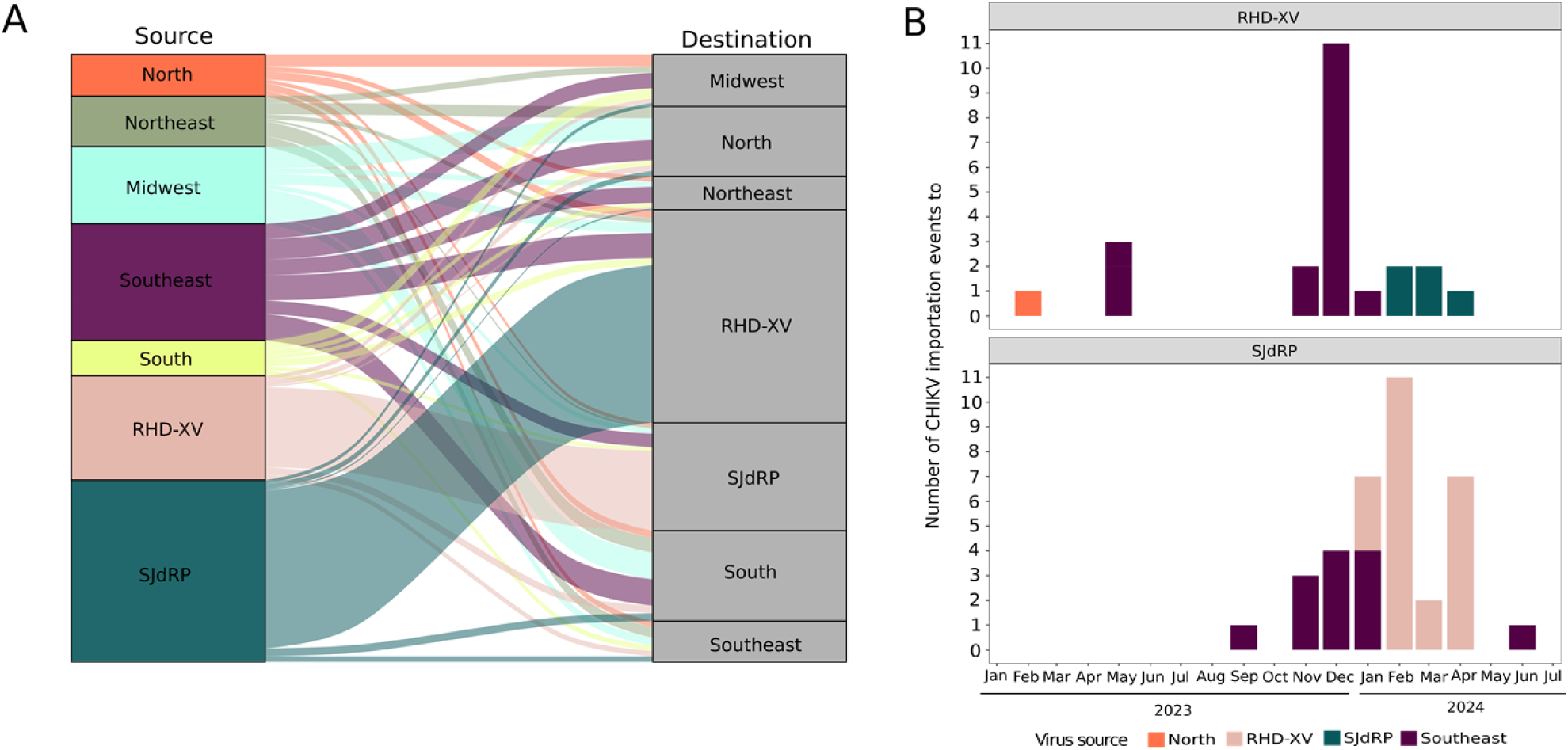
Chikungunya virus exchange across the region. A) Pattern of CHIKV importation and exportation events among SJdRP, RHD-XV municipalities and all Brazilian regions (North, Northeast, Midwest, Southeast, and South). B) Number of sequences imported to RHD-XV municipalities and SJdRP, per month. Colors represent the source of CHIKV virus importations.

By annotating the internal nodes and tips of our phylogenetic tree to geographic locations, we were able to trace CHIKV importation events within the RHD-XV region and SJdRP (Fig. 6B, S6 Table). Our dataset indicated several CHIKV importation events from both the North and Southeast regions to municipalities within the RHD-XV region in early 2023 (Fig 6B). Meanwhile, importation to SJdRP was first recorded in September 2023. Starting in November 2023 a significant increase was observed in CHIKV importation events in both sampled locations (RHD-XV and SJdRP), mainly in December 2023 and February 2024 (Fig. 6B, S6 Table); this increased level of viral exchange aligns with the CHIKV epidemic reflected in the substantial number of infected mosquitoes collected during the same period.

## Discussion

Arboviral diseases pose a significant challenge to public health, particularly in tropical countries with conditions that favor the proliferation and spread of vectors. Brazil is currently endemic for several arboviruses; in 2024 alone, epidemics of DENV (serotypes 1-3), Oropouche (OROV), and CHIKV have been reported [7,10,11,47]. Of these viruses, CHIKV is particularly concerning due to its potential to severely impact quality of life in infected individuals, causing debilitating and chronic symptoms [10]. Accurate differential diagnosis is crucial for better treatment and prognosis of long-term conditions. For this reason, continuous surveillance of acute febrile illness combined with entomological monitoring are important tools for early detection of new viral introductions that could lead to higher numbers of cases.

In this study, we combined epidemiological, entomological, and genomic data to better understand the spatiotemporal dynamics and transmission patterns of CHIKV in northwestern São Paulo state, and found annual epidemic waves been visible since the introduction of this virus into Brazil. Although during 2017–2021 the incidence rate was notably higher in Northeastern states, an increase in cases has been reported from 2022 onward in various regions of Brazil (North, Midwest, and especially the Southeast). These observations are consistent with several studies revealing similar epidemiological patterns across different Brazilian regions [48–52]. In line with our results, transmission hotspots have been identified in Pará and Tocantins in the North, and in Rio de Janeiro and eastern Minas Gerais in the Southeast region [49].

Overall, São Paulo (in the Southeast) has experienced fewer cases of CHIKF compared to other states, which is confirmed by low anti-CHIKV IgG seroprevalence in major cities such as SJdRP and Ribeirão Preto [8,9]. However, our findings indicate that since April 2023 the number of cases in São Paulo has been rising. This was one reason we selected SJdRP, a city without previous CHIKV outbreaks and only cryptic circulation of this virus [9], as a site for active entomological surveillance to detect introductions or highly active arboviruses.

From October 2023 to July 2024 we collected *Aedes* and *Culex* mosquitoes, which exhibited a combined CHIKV positivity rate of 6.67%. This rate of infection in field-captured mosquitoes was higher than previously reported in natural populations of culicids in SJdRP [9] or other regions of Brazil [53,54]. Positivity rose from January to May 2024 in the captured specimens, and most infected mosquitoes were *Ae. aegypti* females. These findings correspond with the rising number of reported CHIKF cases among SJdRP residents, demonstrating a positive relationship between infected vectors and human cases, as expected.

Our study indicates that CHIKV infections exhibit a well-established seasonal pattern similar to dengue epidemics, which are strongly associated with the rainy season in Brazil when higher vector population densities are observed [48,49,52]. This pattern became more pronounced with the decline observed in the number of *Aedes* mosquitoes collected, a decrease in the mosquito positivity rate, and a reduction in human cases starting in June 2024. One factor that may influence decreased CHIKV circulation is less average rainfall in late autumn/early winter in the Southern Hemisphere, which can significantly affect the availability of breeding sites and in turn impact the seasonality of the vector population, as several studies have demonstrated [55,56]. In fact, the São José do Rio Preto municipal water and sewer authority (SEMAE-RP)[15] reported a reduction in rainfall in the city. Temporal shifts in *Aedes* and *Culex* populations (likely resulting from niche competition) may also have contributed to the observed decrease in CHIKV circulation [55,57–61]. We also found a substantial number of male *Aedes* and *Culex* mosquitoes carrying CHIKV: this suggests the presence of vertical and/or sexual transmission of CHIKV, which may contribute to persistent infection and transmission during unfavorable or inter-epidemic periods, as some studies in field-collected mosquitoes have reported [62,63]. Another noteworthy finding is both male and female *Culex* mosquitoes carrying CHIKV, even though *Culex* species are not recognized as competent vectors for this virus. While some studies have detected CHIKV in *Culex* mosquitoes, particularly during epidemics [53,64], and have suggested that these mosquitoes might maintain CHIKV transmission during outbreaks [65], further investigation into the competence of this vector is required. There is currently no conclusive evidence establishing *Culex* mosquitoes as competent vectors for CHIKV transmission.

Furthermore, the presence of CHIKV in naturally infected male mosquitoes in October 2023, even though zero human cases of CHIKF were reported that month, suggests that the virus was already circulating widely in the city well before cases began to climb in November 2023. This observation indicates potential underreporting of CHIKF cases throughout 2023. These findings are supported by data from the SJdRP municipal health department indicating that the majority of confirmed arboviral cases in 2023 and 2024 are attributed to DENV, which contrasts with our results [16]. According to the Brazilian Ministry of Health’s Epidemiological Bulletin [10] there is a noted discrepancy between reported cases of dengue and chikungunya across several Brazilian states: dengue is the predominant diagnosis. This highlights the critical need for differential diagnosis using laboratory assays like RT-qPCR to identify additional cases of CHIKF, and suggests that CHIKV may be underreported in certain regions [10].

Our spatiotemporal analysis of CHIKV in northwestern São Paulo state revealed multiple introduction events, primarily originating from the Southeast (including Minas Gerais and other cities in São Paulo state) and the North. These introductions were crucial for the establishment and dissemination of the virus, and marked the first outbreak of CHIKV in this area. Factors contributing to this initial epidemic include a high density of competent vectors and a substantial susceptible population for CHIKV infections [6,9,66]. Similarly, Souza et al. [48] identified the introduction of a new lineage of the CHIKV-ECSA variant, closely related to CHIKV sequences circulating in São Paulo during 2021–2022, as the cause of CHIKV recurrence in Ceará and Tocantins (Northeast and North regions, respectively) in 2022. This resurgence predominantly affected cities that had reported few or no CHIKV cases in previous epidemic waves. Furthermore, while the Northeast region of Brazil has been implicated as the original source of CHIKV in the country [50,67], the recent establishment of CHIKV in Southeast Brazil has significantly facilitated exchange of this virus throughout the country, as our migration pattern analysis shows.

Our results also indicate that the introduction of CHIKV in the Northwest region of São Paulo likely occurred in early 2023, and that the virus circulated for several months prior to the increase in human cases observed in November 2023. This observation supports our findings of both male and female mosquitoes carrying CHIKV, and underscores the urgent need to implement an entomological surveillance program for early detection of circulating arboviruses and anticipate future outbreaks.

## Conclusion

Implementation of an entomological surveillance system successfully detected the spread of CHIKV across SJdRP, revealing an infection rate of 6.67%. Our findings suggest that the virus was extensively disseminated within the natural mosquito population, potentially through vertical or sexual transmission, which might contribute to sustained infection and transmission during periods of unfavorable conditions or inter-epidemic intervals. We also observed a monthly increase in the positivity rate among the vector population from December to April 2024, which correlates with the rising number of CHIKF cases reported in the city during the same period. The resurgence of CHIKV in northwestern São Paulo state is the result of several introduction events (mainly from the Southeast and North of the country) that helped establish the virus in the highly dense vector population and, in turn, its extensive spread, leading to a major CHIKF epidemic in this geographical area. Given the cocirculation of multiple arboviruses in several municipalities within São Paulo state, entomological surveillance and ongoing monitoring of patient samples are crucial for health authorities to implement more effective control interventions in order to interrupt transmission cycles in endemic regions or mitigate new epidemic waves.

## Data availability

All the chikungunya genomes generated and analyzed in this study are available in the GenBank NCBI and GISAID databases, under accession numbers provided in the S5 Table. Further information about the generated genomes such as collection date, location and host are provided in Table S5. All data used for epidemiological and phylogenetic analyses are available in the Mendeley Data repository (10.17632/6mtrbck5sj.1)

## Acknowledgments

We wish to thank all our colleagues at the Hospital de Base de São José do Rio Preto for the support they provided during sample collection. We are also grateful to the Multiuser Laboratory (LMU) at the São José do Rio Preto School of Medicine (FAMERP) and the Instituto de Biotecnologia at São Paulo State University (UNESP) in Botucatu, Brazil for allowing us to use the Illumina MiSeq system.

## Author contributions

CAB, MCPP and MLN conceived and designed the study. CAB, MCPP, OBN, GPM, MVMF, APL, RMTM and JTD collected and identified mosquito samples. CAB, MCPP, OBN, MVMF and APL processed and screened mosquito samples. MLB collected and screened human samples. CAB, MCPP, OBN, MVMF and APL, curated the metadata and performed molecular screening in mosquito samples. AFN provided metadata from SJdRP. CFE curated metadata for the human samples. LS standardized the arbovirus sequencing protocol. CAB and BCM performed sequencing. CAB performed epidemiological, geoprocessing and phylogenetic analyses. CAB and MCPP interpreted the data. CAB, MCPP, BCM and CFE wrote the first draft of the manuscript. CAB, MCPP, BCM, CFE, NV and MLN edited and revised the manuscript. NV and MLN provided the resources for the survey. All authors approved the final version of this manuscript.

## Ethics statement

This study was approved by the Ethics Committee of the São José do Rio Preto School of Medicine (FAMERP), process CAAE 79090324.8.0000.5415 on April 30, 2024, and CAAE 02078812.8.0000.5415, on July 03, 2012 amendment on May 11, 2016.

## Supporting information

**S1 Table.** Laboratory confirmed cases of chikungunya fever retrieved from the National Reportable Disease Information System (SINAN) for all Brazilian states, January 2017–July 2024.

**S2 Table.** Incidence of chikungunya fever per 100,000 inhabitants, January 2017–July 2024, for all Brazilian states.

**S3 Table.** Chikungunya virus screening and genome sequencing information for collected mosquitoes, October 2023–July 2024. Mosquitoes were collected in six neighborhoods in São José do Rio Preto, São Paulo, Brazil, and classified according to species and sex.

**S4 Table.** Cycle threshold values observed in chikungunya-positive mosquito samples.

**S5 Table.** Chikungunya genomes used in the phylogenetic analysis. The dataset is composed of 782 Brazilian complete genomes (149 from this study) from all Brazilian regions and from different hosts, collected from 2014 to 2024.

**S6 Table.** Chikungunya virus exchange events among São José do Rio Preto, the cities within the Regional Health Department XV region and all Brazilian regions (North, Northeast, Midwest, Southeast and South).

**S1 Fig. Chikungunya virus molecular testing.** Cycle threshold range obtained for CHIKV positive samples of *Aedes* and *Culex* mosquitoes.

## References

1. Peter M. Howley MD DMKSW. Fields Virology: Emerging Viruses. 7th ed. 2020.

2. Laporta GZ, Potter AM, Oliveira JFA, Bourke BP, Pecor DB, Linton YM. Global Distribution of Aedes aegypti and Aedes albopictus in a Climate Change Scenario of Regional Rivalry. Insects. 2023;14. doi:10.3390/insects14010049

3. Leparc-Goffart I, Nougairede A, Cassadou S, Prat C, De Lamballerie X. Chikungunya in the Americas. The Lancet. 2014. doi:10.1016/S0140-6736(14)60185-9

4. Nunes MRT, Faria NR, de Vasconcelos JM, Golding N, Kraemer MUG, de Oliveira LF, et al. Emergence and potential for spread of Chikungunya virus in Brazil. BMC Med. 2015;13. doi:10.1186/s12916-015-0348-x

5. Ministério da Saúde. DATASUS. In: https://datasus.saude.gov.br/ [Internet]. 2024 [cited 17 Oct 2024]. Available: http://tabnet.datasus.gov.br/cgi/tabcgi.exe?sinannet/cnv/chikunbr.def

6. Fonseca Júnior DP da, Serpa LLN, Barbosa GL, Pereira M, Holcmam MM, Voltolini JC, et al. Vectors of arboviruses in the state of São Paulo: 30 years of Aedes aegypti and Aedes albopictus. Rev Saude Publica. 2019;53. doi:10.11606/s1518-8787.2019053001264

7. Siqueira TS, Silva LS, de Holanda JRC, Carvalho SCC, Silva JRS, Santos VS. Spatial clustering of dengue cases during the 2024 epidemic in Brazil. J Travel Med. 2024. doi:10.1093/jtm/taae093

8. Slavov SN, Otaguiri KK, Bianquini ML, Bitencourt HTO, Chagas MCM, Guerreiro DS de S, et al. Seroprevalence of Chikungunya virus in blood donors from Northern and Southeastern Brazil. Hematol Transfus Cell Ther. 2018;40. doi:10.1016/j.htct.2018.04.002

9. Zini N, Ávila MHT, Cezarotti NM, Parra MCP, Banho CA, Sacchetto L, et al. Cryptic circulation of chikungunya virus in São Jose do Rio Preto, Brazil, 2015–2019. PLoS Negl Trop Dis. 2024;2024. doi:10.1371/journal.pntd.0012013

10. MINISTÉRIO DA SAÚDE SDVESEA. Monitoramento das arboviroses e balanço de encerramento do Comitê de Operações de Emergência (COE) Dengue e outras Arboviroses 2024 MINISTÉRIO DA SAÚDE. 2024. Available: file:///C:/Users/User/Downloads/Boletim%20Epidemiol%C3%B3gico%20-%20Volume%2055%20-%20n%C2%BA%2011%20(1).pdf

11. Adelino T, Lima M, Guimarães NR, Xavier J, Fonseca V, Tomé LMR, et al. Resurgence of Dengue Virus Serotype 3 in Minas Gerais, Brazil: A Case Report. Pathogens. 2024;13. doi:10.3390/pathogens13030202

12. Fujita DM, Salvador FS, da Silva Nali LH, de Andrade Júnior HF. Silent spread of DENV-3 in Brazil: autochthonous outbreak in São Paulo after 15 years. Journal of Travel Medicine. 2024. doi:10.1093/jtm/taad166

13. da Costa VG, Saivish M V., Sinhorini PF, Nogueira ML, Rahal P. A meta-analysis of Chikungunya virus in neurological disorders. Infectious Diseases Now. Elsevier Masson s.r.l.; 2024. doi:10.1016/j.idnow.2024.104938

14. IBGE Instituto Brasileiro de Geografia e Estatística. CENSO 2022. In: https://censo2022.ibge.gov.br/panorama/ [Internet]. [cited 17 Oct 2024]. Available: https://censo2022.ibge.gov.br/panorama/

15. Serviço Municipal Autônomo de Água e Esgoto de São José do Rio Preto. Índice Pluviométrico. In: https://semae.riopreto.sp.gov.br/indice-pluviometrico/ [Internet]. 2024 [cited 17 Oct 2024]. Available: https://semae.riopreto.sp.gov.br/indice-pluviometrico/

16. Secretaria da Saúde - Prefeitura de São José do Rio Preto. Vigilância Epidemiológica - Boletins Epidemiológicos Dengue 2024. In: https://saude.riopreto.sp.gov.br/transparencia/boletim_dengue_saude_riopreto.php [Internet]. 2024 [cited 17 Oct 2024]. Available: https://saude.riopreto.sp.gov.br/transparencia/boletim_dengue_saude_riopreto.php

17. Rotraut A. G. B. Consoli and Ricardo Lourenço de Oliveira. Principais mosquitos de importância sanitária no Brasil. Editora FIOCRUZ. Rio de Janeiro: Editora FIOCRUZ; 1994.

18. Oswaldo Paulo Forattini. Culicidologia Médica: Princípios Gerais, Morfologia, Glossário Taxonômico. 1st ed. 1996.

19. Instituto Brasileiro de Geografia e Estatística (IBGE). Malha Municipal. In: https://www.ibge.gov.br/geociencias/organizacao-do-territorio/malhas-territoriais/15774-malhas.html [Internet]. 2024 [cited 17 Oct 2024]. Available: https://www.ibge.gov.br/geociencias/organizacao-do-territorio/malhas-territoriais/15774-malhas.html

20. RStudio Team. RStudio: Integrated Development for R. RStudio, Inc., Boston, MA. 2021.

21. Pebesma E. Simple features for R: Standardized support for spatial vector data. R Journal. 2018;10. doi:10.32614/rj-2018-009

22. Pebesma E, Bivand R. Spatial Data Science: With Applications in R. Spatial Data Science: With Applications in R. 2023. doi:10.1201/9780429459016

23. Wilkinson L. ggplot2: Elegant Graphics for Data Analysis by WICKHAM, H. Biometrics. 2011;67. doi:10.1111/j.1541-0420.2011.01616.x

24. Instituto Brasileiro de Geografia e Estatística IBGE. SIDRA - Banco de Tabelas Estatísticas. In: https://sidra.ibge.gov.br/home/lspa/brasil. 18 Oct 2024.

25. Secretaria da Saúde - Prefeitura de São José do Rio Preto. Vigilância Epidemiológica - Boletins Epidemiológicos Chikungunya. In: https://saude.riopreto.sp.gov.br/transparencia/boletim_chikungunya_saude_riopreto.php [Internet]. 2024 [cited 17 Oct 2024]. Available: https://saude.riopreto.sp.gov.br/transparencia/boletim_chikungunya_saude_riopreto.php

26. MacHado DC, Mondini A, Dos Santos Santana V, Yonamine PTK, Chiaravalloti Neto F, De Andrade Zanotto PM, et al. First identification of culex flavivirus (flaviviridae) in brazil. Intervirology. 2012;55. doi:10.1159/000337166

27. Lanciotti RS, Kosoy OL, Laven JJ, Panella AJ, Velez JO, Lambert AJ, et al. Chikungunya virus in US travelers returning from India, 2006. Emerg Infect Dis. 2007;13. doi:10.3201/eid1305.070015

28. Simon Andrews. Babraham Bioinformatics - FastQC A Quality Control tool for High Throughput Sequence Data. Soil. 2020;5.

29. Martin M. Cutadapt removes adapter sequences from high-throughput sequencing reads. EMBnet J. 2011;17. doi:10.14806/ej.17.1.200

30. Langmead B, Salzberg SL. Fast gapped-read alignment with Bowtie 2. Nat Methods. 2012;9. doi:10.1038/nmeth.1923

31. Li H, Handsaker B, Wysoker A, Fennell T, Ruan J, Homer N, et al. The Sequence Alignment/Map format and SAMtools. Bioinformatics. 2009;25. doi:10.1093/bioinformatics/btp352

32. Li H, Durbin R. Fast and accurate short read alignment with Burrows-Wheeler transform. Bioinformatics. 2009;25. doi:10.1093/bioinformatics/btp324

33. Castellano S, Cestari F, Faglioni G, Tenedini E, Marino M, Artuso L, et al. Ivar, an interpretation-oriented tool to manage the update and revision of variant annotation and classification. Genes (Basel). 2021;12. doi:10.3390/genes12030384

34. Vilsker M, Moosa Y, Nooij S, Fonseca V, Ghysens Y, Dumon K, et al. Genome Detective: An automated system for virus identification from high-throughput sequencing data. Bioinformatics. 2019;35. doi:10.1093/bioinformatics/bty695

35. Elbe S, Buckland-Merrett G. Data, disease and diplomacy: GISAID’s innovative contribution to global health. Global Challenges. 2017;1. doi:10.1002/gch2.1018

36. Clark K, Karsch-Mizrachi I, Lipman DJ, Ostell J, Sayers EW. GenBank. Nucleic Acids Res. 2016;44. doi:10.1093/nar/gkv1276

37. Katoh K, Standley DM. MAFFT multiple sequence alignment software version 7: Improvements in performance and usability. Mol Biol Evol. 2013;30. doi:10.1093/molbev/mst010

38. Larsson A. AliView: A fast and lightweight alignment viewer and editor for large datasets. Bioinformatics. 2014;30. doi:10.1093/bioinformatics/btu531

39. Nguyen LT, Schmidt HA, Von Haeseler A, Minh BQ. IQ-TREE: A fast and effective stochastic algorithm for estimating maximum-likelihood phylogenies. Mol Biol Evol. 2015;32. doi:10.1093/molbev/msu300

40. Kalyaanamoorthy S, Minh BQ, Wong TKF, Von Haeseler A, Jermiin LS. ModelFinder: Fast model selection for accurate phylogenetic estimates. Nat Methods. 2017;14. doi:10.1038/nmeth.4285

41. Minh BQ, Nguyen MAT, Von Haeseler A. Ultrafast approximation for phylogenetic bootstrap. Mol Biol Evol. 2013;30. doi:10.1093/molbev/mst024

42. Anisimova M, Gascuel O. Approximate likelihood-ratio test for branches: A fast, accurate, and powerful alternative. Syst Biol. 2006;55. doi:10.1080/10635150600755453

43. Xu S, Li L, Luo X, Chen M, Tang W, Zhan L, et al. Ggtree: A serialized data object for visualization of a phylogenetic tree and annotation data. iMeta. 2022;1. doi:10.1002/imt2.56

44. Rambaut A, Lam TT, Carvalho LM, Pybus OG. Exploring the temporal structure of heterochronous sequences using TempEst (formerly Path-O-Gen). Virus Evol. 2016;2. doi:10.1093/ve/vew007

45. Sagulenko P, Puller V, Neher RA. TreeTime: Maximum-likelihood phylodynamic analysis. Virus Evol. 2018;4. doi:10.1093/ve/vex042

46. Banho CA, de Carvalho Marques B, Sacchetto L, Lima AKS, Parra MCP, Lima ARJ, et al. Dynamic clade transitions and the influence of vaccination on the spatiotemporal circulation of SARS-CoV-2 variants. NPJ Vaccines. 2024;9. doi:10.1038/s41541-024-00933-w

47. Scachetti GC, Forato J, Claro IM, Hua X, Salgado BB, Vieira A, et al. Reemergence of Oropouche virus between 2023 and 2024 in Brazil. medRxiv. 2024. doi:10.1101/2024.07.27.24310296

48. de Souza WM, de Lima STS, Simões Mello LM, Candido DS, Buss L, Whittaker C, et al. Spatiotemporal dynamics and recurrence of chikungunya virus in Brazil: an epidemiological study. Lancet Microbe. 2023;4. doi:10.1016/S2666-5247(23)00033-2

49. Yakob L. Predictable Chikungunya Infection Dynamics in Brazil. Viruses. 2022;14. doi:10.3390/v14091889

50. Xavier J, Alcantara LCJ, Fonseca V, Lima M, Castro E, Fritsch H, et al. Increased interregional virus exchange and nucleotide diversity outline the expansion of chikungunya virus in Brazil. Nat Commun. 2023;14. doi:10.1038/s41467-023-40099-y

51. Silva A do C, Silva A do C, de Castro PASV, Ávila IR, Bezerra JMT. Prevalence and epidemiological aspects of Chikungunya fever in states of the Northeast region of Brazil: A systematic review. Acta Tropica. 2023. doi:10.1016/j.actatropica.2023.106872

52. Vidal ERN, Frutuoso LCV, Duarte EC, Peixoto HM. Epidemiological burden of Chikungunya fever in Brazil, 2016 and 2017. Tropical Medicine and International Health. 2022;27. doi:10.1111/tmi.13711

53. Cruz ACR, Neto JPN, da Silva SP, da Silva EVP, Pereira GJG, Santos MM, et al. Chikungunya virus detection in aedes aegypti and culex quinquefasciatus during an outbreak in the amazon region. Viruses. 2020;12. doi:10.3390/v12080853

54. Aragão CF, Pinheiro VCS, Neto JPN, Da Silva EVP, Pereira GJG, Do Nascimento BLS, et al. Natural infection of Aedes aegypti by Chikungunya and Dengue type 2 Virus in a transition area of north-northeast Brazil. Viruses. 2019;11. doi:10.3390/v11121126

55. Heinisch MRS, Diaz-Quijano FA, Chiaravalloti-Neto F, Menezes Pancetti FG, Rocha Coelho R, dos Santos Andrade P, et al. Seasonal and spatial distribution of Aedes aegypti and Aedes albopictus in a municipal urban park in São Paulo, SP, Brazil. Acta Trop. 2019;189. doi:10.1016/j.actatropica.2018.09.011

56. Dibo MR, Chierotti AP, Ferrari MS, Mendonça AL, Neto FC. Study of the relationship between Aedes (Stegomyia) aegypti egg and adult densities, dengue fever and climate in Mirassol, state of São Paulo, Brazil. Mem Inst Oswaldo Cruz. 2008;103. doi:10.1590/S0074-02762008000600008

57. Barbosa RMR, Regis LN. Monitoring temporal fluctuations of Culex quinquefasciatus using oviposition traps containing attractant and larvicide in an urban environment in recife, Brazil. Mem Inst Oswaldo Cruz. 2011;106. doi:10.1590/S0074-02762011000400011

58. Medeiros AS, Marcondes CB, De Azevedo PRM, Jerônimo SMR, Silva VPMEE, De De Ximenes MFFM. Seasonal variation of potential flavivirus vectors in an urban biological reserve in Northeastern Brazil. J Med Entomol. 2009;46. doi:10.1603/033.046.0630

59. Costa IMP, Calado DC. Incidência dos casos de dengue (2007-2013) e distribuição sazonal de culicídeos (2012-2013) em Barreiras, Bahia. Epidemiol Serv Saude. 2016;25. doi:10.5123/S1679-49742016000400007

60. Degener CM, de Ázara TMF, Roque RA, Codeço CT, Nobre AA, Ohly JJ, et al. Temporal abundance of Aedes aegypti in Manaus, Brazil, measured by two trap types for adult mosquitoes. Mem Inst Oswaldo Cruz. 2014;109. doi:10.1590/0074-0276140234

61. Marini G, Guzzetta G, Baldacchino F, Arnoldi D, Montarsi F, Capelli G, et al. The effect of interspecific competition on the temporal dynamics of Aedes albopictus and Culex pipiens. Parasit Vectors. 2017;10. doi:10.1186/s13071-017-2041-8

62. Jain J, Kushwah RBS, Singh SS, Sharma A, Adak T, Singh OP, et al. Evidence for natural vertical transmission of chikungunya viruses in field populations of Aedes aegypti in Delhi and Haryana states in India—a preliminary report. Acta Trop. 2016;162. doi:10.1016/j.actatropica.2016.06.004

63. Dzul-Manzanilla F, Martínez NE, Cruz-Nolasco M, Gutiérrez-Castro C, López-Damián L, Ibarra-López J, et al. Evidence of vertical transmission and co-circulation of chikungunya and dengue viruses in field populations of Aedes aegypti (L.) from Guerrero, Mexico. Trans R Soc Trop Med Hyg. 2016;110. doi:10.1093/trstmh/trv106

64. da Silva Neves NA, da Silva Ferreira R, Morais DO, Pavon JAR, de Pinho JB, Slhessarenko RD. Chikungunya, Zika, Mayaro, and Equine Encephalitis virus detection in adult Culicinae from South Central Mato Grosso, Brazil, during the rainy season of 2018. Brazilian Journal of Microbiology. 2022;53. doi:10.1007/s42770-021-00646-5

65. Lutomiah J, Mulwa F, Mutisya J, Koskei E, Langat S, Nyunja A, et al. Probable contribution of Culex quinquefasciatus mosquitoes to the circulation of chikungunya virus during an outbreak in Mombasa County, Kenya, 2017–2018. Parasit Vectors. 2021;14. doi:10.1186/s13071-021-04632-6

66. Parra MCP, Lorenz C, Dibo MR, de Aguiar Milhim BHG, Guirado MM, Nogueira ML, et al. Association between densities of adult and immature stages of Aedes aegypti mosquitoes in space and time: implications for vector surveillance. Parasit Vectors. 2022;15. doi:10.1186/s13071-022-05244-4

67. Souza UJB de, Santos RN dos, Giovanetti M, Alcantara LCJ, Galvão JD, Cardoso FDP, et al. Genomic Epidemiology Reveals the Circulation of the Chikungunya Virus East/Central/South African Lineage in Tocantins State, North Brazil. Viruses. 2022;14. doi:10.3390/v14102311

